# *‘I had no life. I was only existing’*. Factors shaping the mental health and wellbeing of people experiencing long Covid: a qualitative study

**DOI:** 10.1101/2021.10.13.21264855

**Authors:** Alexandra Burton, Henry Aughterson, Daisy Fancourt, Keir EJ Philip

**Affiliations:** Department of Behavioural Science and Health, University College London, United Kingdom; National Heart and Lung Institute, Imperial College London, London, United Kingdom; NIHR Imperial Biomedical Research Centre, London, United Kingdom

## Abstract

**Background:** Around one in 10 people who have COVID-19 report persistent symptoms or ‘long Covid’. Impaired mental health and wellbeing is commonly reported including anxiety, depression and reduced quality of life. There is however, limited in-depth research exploring *why* mental health and wellbeing have been impacted among people experiencing long Covid.

**Aims:** To explore factors impacting mental health and wellbeing, from the perspective of people with long Covid.

**Method:** Semi-structured qualitative interviews that were audio-recorded and transcribed. Data were analysed using reflexive thematic analysis. 21 people with long Covid participated in the study. Participants were eligible to take part if they self-reported a positive swab test/antibody test, or one or more commonly reported COVID-19 symptoms at illness onset and experiences of one or more long Covid symptom three or more weeks following illness onset.

**Results:** Five themes were identified across participant accounts regarding factors impacting mental health and wellbeing including i) experiences of care and understanding from others; ii) lack of service and treatment options; iii) severe disruption to daily life iv) uncertainty of illness trajectories and v) changes to identity.

**Conclusions:** People with long Covid experience a range of factors that negatively impact their mental health and wellbeing. Providing patient centred health services that integrate the rapidly evolving research in this area is important, as are peer support groups and supported approaches to self-management.

## Introduction

The COVID-19 pandemic, caused by Severe Acute Respiratory Syndrome Coronavirus-2 (SARS-CoV-2), has had major consequences for health and wellbeing globally. The impact of SARS-CoV-2 infection varies widely between individuals from being asymptomatic to multi-organ failure and death. For many people, symptoms persist far beyond the acute phase of the disease^1-4^. Various names have been given to this condition including ongoing symptomatic COVID-19, post-acute COVID-19 syndrome, ‘long-haulers’, and long Covid^1,5^. Here we use the term long Covid, as it has become popular among many people with the condition and is widely used in the medical literature^1^, however we appreciate that discussions continue^1,5,6^.

Long Covid prevalence estimates are inconsistent due to the rapidly evolving research base, non-standardised diagnostic criteria, and different populations studied^7^. Current research suggests approximately 10% of people will have ongoing symptoms 12 weeks after initial infection, and substantial numbers have symptoms lasting over six months^1 3^. Given the large global burden of infections with SARS-CoV-2, even a small proportion of people with persistent symptoms represents a major public health issue. Long Covid symptoms can be variable and wide-ranging, relating to multiple organ systems including respiratory, cardiovascular, neurological, psychological/psychiatric, gastrointestinal, musculoskeletal and dermatological; or they can be general and non-specific^8^. Symptoms are likely to result from multiple potentially overlapping disease mechanisms^1 9^. The most commonly reported symptoms include fatigue, breathlessness, cardiovascular abnormalities, decreased cognitive function, difficulties sleeping, and abnormalities in taste and smell^10^. Comparisons have been made between long Covid and other conditions including post-intensive care syndrome, myalgic-encephalitis/chronic fatigue syndrome (ME/CFS), and the sequalae of infection with coronaviruses like SARS and MERS which can cause protracted multisystem disease^1 11^. However, although certain shared features exist, caution should be applied when drawing parallels with symptoms experienced in other conditions, as doing so may detract from appropriate investigation and management^6^.

An evolving research base has sought to understand the types of services and support available to people experiencing long Covid. Reports suggest multiple difficulties accessing appropriate care including uncertainty about treatment options and a lack of coordinated care^12 13^. These experiences have resulted in patients resorting to self-management of symptoms through trial and error and via information and advice taken from informal peer support networks^14^.

Mental health and wellbeing impacts are commonly reported by people with long Covid, in particular features of anxiety, depression and post-traumatic stress disorder (PTSD) ^8 9 15-17^, with cognitive processing disorders, or ‘brain-fog’, also frequently reported^8 9^. Mental health impacts are likely multifactorial in origin, including the psychosocial context of the pandemic^18-20^; pre-existing individual susceptibilities^18 21^; and physical biological processes related to SARS-CoV-2 infection^19 21 22^. Although mental health impacts are widely reported, there is limited in-depth research exploring the perspectives of people with long Covid regarding factors shaping their mental health and wellbeing. Addressing this gap has been identified as a key priority by an international, multi-stakeholder forum^23^, to improve understanding, promote empathic healthcare interactions, and facilitate development of patient centred and appropriate healthcare provision for people with long Covid. Therefore, this study aimed to explore the perspectives of people with long Covid regarding how their mental health and wellbeing has been affected.

## Methods

### Sample and recruitment

We conducted semi-structured qualitative interviews with people with self-reported long Covid. Participants were recruited via an online long Covid support group, an advertisement in the UCL Covid Social Study^24^ newsletter and through social media. 74 people responded to the invitation to take part and from this participant pool, 53 people were purposively sampled to ensure that people of different ages, ethnicity and genders were represented in the study. All 53 potential participants were sent a study information sheet and a screening form to determine eligibility. 29 screening forms were returned with no further response from 23 people and 1 person declining further participation. 21 people went on take part in the study.

To assess participant eligibility for the study we used the post-acute COVID-19 criteria proposed by Greenhalgh et al 2020, which was broadly representative of the consensus at the time^25^. Eligibility criteria consisted of 1) a positive swab test/antibody test OR 1+ commonly reported symptom at illness onset (persistent cough, loss or change in taste/smell, high temperature) AND 2) experiencing one or more broader symptoms three or more weeks following the onset of their first symptoms^25^ (S1 Appendix). Eligible participants were encouraged to ask questions about the study before providing written informed consent to take part and completing a demographics form. Ethical approval to conduct the study was received from the UCL Ethics Committee (Project ID 14895/005).

### Data collection

All interviews followed a semi-structured topic guide, and were audio recorded. Questions explored i) onset and initial impact of COVID-19 symptoms, ii) ongoing impact and development of longer-term health problems, iii) impact of long Covid on social lives and mental health, and iv) worries about the future. Example topic guide questions are listed in Figure 2 and the full topic guide is provided as supplementary material (S2 Appendix). Interviews were conducted by a female postdoctoral applied mental health services researcher (AB) via video (14 interviews) or telephone call (7 interviews) depending on participant preference. Interviews lasted on average 67 minutes (range 41-99 minutes) and participants received a £ 10 shopping voucher for their time. Data collection continued until no new concepts related to mental health and wellbeing were discussed by interviewees^26^.

**Figure 2. Topic guide question examples**

- *Can you tell me about when you first suspected you had COVID-19? What was the impact on normal life at the time?*
- *Can you describe the ongoing impact and development of longer-term health problems?*
- *Has your experience of COVID-19/long COVID had any impact on your social life?*
- *How do you feel about the changes that have been brought about by COVID-19? Have they had any impact on your mental health or wellbeing?*
- *Have you been doing/planning anything to help with this?*
- *Has the pandemic/long Covid meant that you have any worries for the future?*

### Data analysis

Audio files were transcribed verbatim by an external transcription company. Personal and identifiable data were removed from transcripts to maintain confidentiality. Transcripts were then imported into NVIVO12 software and analysed using reflexive thematic analysis^27^. Analysis was inductive whereby codes were generated from the text using line by line coding. Three transcripts (14%) were independently double coded by three researchers (AB, HA (a medical trainee and doctoral student in behavioural science and health) and KEJP (a physician and clinical research fellow with training in qualitative research)) who met to discuss emerging codes and themes and generate a coding framework. HA then applied the coding framework to all remaining transcripts, adding new codes as transcripts became available and until no new codes were identified. Codes were then arranged into themes pertaining to the research questions. The research team met to finalise themes and approve the final report.

## Results

We interviewed 21 participants between November 2020 and September 2021. Participants were aged between 26-70 years old (mean = 47), predominantly female (67%), and White British (67%). All participants reported a case of suspected or confirmed COVID-19 between 28^th^ February and 11^th^ January 2021 and experienced on average 12 symptoms (range 4-18 symptoms) for 29 weeks at the time of interview (range 8-52 weeks), with the most common symptoms being fatigue (95%), muscle pain/weakness (86%), shortness of breath (81%), difficulties concentrating (76%) and memory lapses (71%). 52% of participants reported a confirmed positive swab test or had tested positive for antibodies. Four participants were hospitalised during the initial phase of their illness. All participants were still experiencing symptoms at the time of the interview. See Table 1 for participant characteristics and Table 2 for symptoms and self-reported laboratory confirmation of SARS-CoV-2 infection.

**Table 1:** Participant characteristics*.

| Characteristics | N = 21 (100%) |
| --- | --- |
| **Age (Range/Mean)** | 26-70 (47) |
| **Female** | 14 (67%) |
| **Ethnicity (self-reported)** |  |
| White British | 14 (67%) |
| Asian (Indian/Pakistani) | 5 (24%) |
| White Irish | 1 (5%) |
| Other Asian Background | 1 (5%) |
| **Marital Status** |  |
| Married/civil partnership/living with partner | 13 (62%) |
| Single | 5 (24%) |
| Divorced/separated | 2 (10%) |
| With partner but not living together | 1 (5%) |
| **Living situation** |  |
| With others | 16 (62%) |
| Alone | 5 (24%) |
| **Employment** |  |
| Employed | 18 (86%) |
| Retired | 2 (10%) |
| Unable to work due to illness | 1 (5%) |
| **Pre-covid physical health problem#** | 10 (48%) |
| **Pre-covid mental health problems+** | 2 (10%) |

\*Data are self-reported using a demographics form.
#Hypothyroid, irritable bowel syndrome, arthritis, asthma, type 2 diabetes, heart failure, hypertension, vestibular migraine, gout, prostate problems, radicular pain, bile acid malabsorption

**Table 2:**
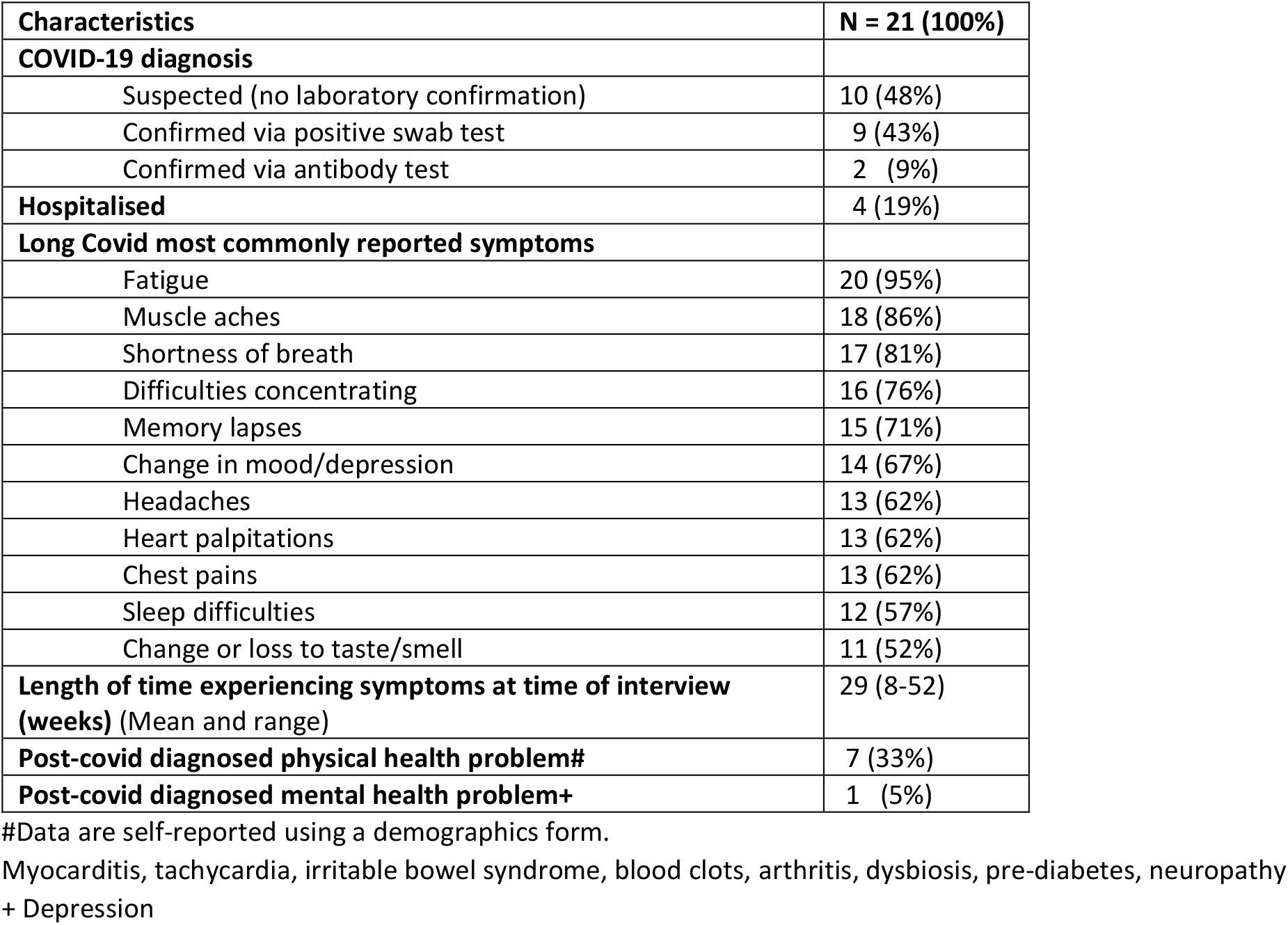
COVID-19 status and symptoms.

| Characteristics | N = 21 (100%) |
| --- | --- |
| **COVID-19 diagnosis** |  |
| Suspected (no laboratory confirmation) | 10 (48%) |
| Confirmed via positive swab test | 9 (43%) |
| Confirmed via antibody test | 2 (9%) |
| **Hospitalised** | 4 (19%) |
| **Long Covid most commonly reported symptoms** |  |
| Fatigue | 20 (95%) |
| Muscle aches | 18 (86%) |
| Shortness of breath | 17 (81%) |
| Difficulties concentrating | 16 (76%) |
| Memory lapses | 15 (71%) |
| Change in mood/depression | 14 (67%) |
| Headaches | 13 (62%) |
| Heart palpitations | 13 (62%) |
| Chest pains | 13 (62%) |
| Sleep difficulties | 12 (57%) |
| Change or loss to taste/smell | 11 (52%) |
| **Length of time experiencing symptoms at time of interview (weeks) (Mean and range)** | 29 (8-52) |
| **Post-covid diagnosed physical health problem#** | 7 (33%) |
| **Post-covid diagnosed mental health problem+** | 1 (5%) |

#Data are self-reported using a demographics form.

### Themes

We identified five themes regarding factors shaping mental health and wellbeing for people living with long Covid: i) care and understanding from others; ii) lack of service and treatment options; iii) severe disruption to daily life; iv) uncertainty of illness trajectories and v) changes to identity. See Table 3 for a list of superordinate themes and sub themes.

**Table 3.**
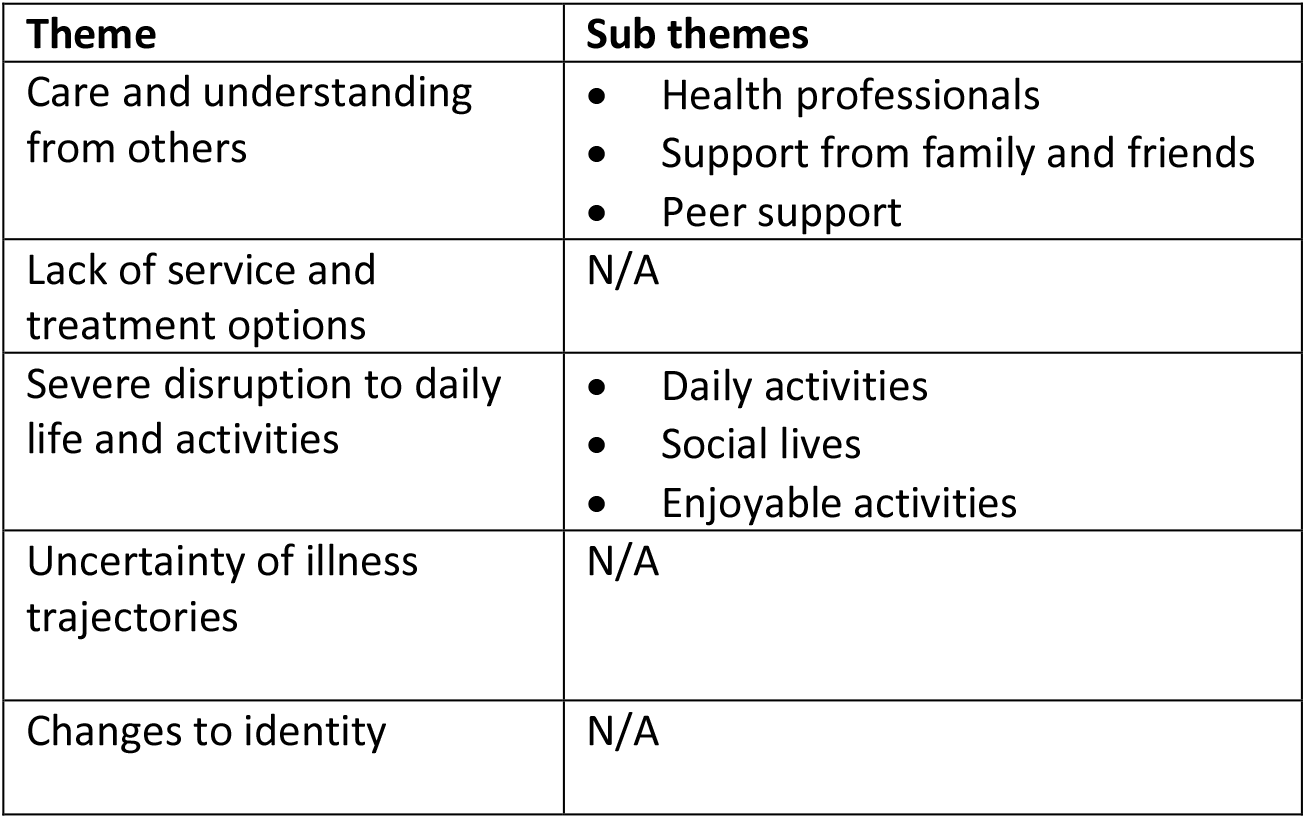
Themes and corresponding sub themes

| Theme | Sub themes |
| --- | --- |
| Care and understanding from others | • Health professionals<br>• Support from family and friends<br>• Peer support |
| Lack of service and treatment options | N/A |
| Severe disruption to daily life and activities | • Daily activities<br>• Social lives<br>• Enjoyable activities |
| Uncertainty of illness trajectories | N/A |
| Changes to identity | N/A |

#### Theme 1: Care and understanding from others

Having a supportive and understanding social network, including friends, family and health professionals was described as important for participant wellbeing, particularly when faced with fluctuating symptoms and uncertainty (Theme 3). When this support was not available, participants described feeling anxious and alone.

#### Health professionals

Participants frequently reported feelings of frustration or anxiety due to a lack of understanding and support from health professionals. Experiences that exacerbated poor mental health included feeling ‘*ignored’, ‘abandoned’* or *‘being brushed aside’*, feeling that health professionals were unwilling to help or listen, a lack of continuity of care, and instances of diagnostic overshadowing whereby symptoms of long Covid were attributed to the participant’s mental health.

> *‘I have come across a few doctors where I’ve felt they’ve made me feel a bit stupid when I’m worrying about certain things*…*So yes, there’s that worrying that people just think you’re making it up. I think it’s just that anxiety thing, for me that I don’t want to be an anxious person. And I think if I sense that I’m talking to a doctor, or sometimes they’ve said it outright, you’re an anxious person, and I think oh great they just think I’m anxious*.*’* (558_female_aged_35-39)

Even when health professionals were unable to offer appropriate medical treatments or answers, it was still important for participants to feel heard and for their stories to be believed and thus validated.

> *‘In the early days, my GP was fantastic… I sent him a letter to tell him what was going on in my life, all my symptoms and everything. And I said, I am so sorry about harassing you. And he phoned me up, and he said, keep harassing me, he said, if you don’t tell me, I don’t know*.*’* (564_female_aged_70-74)

#### Support from family and friends

Participants described the importance of receiving support and empathy from their social networks to protect their wellbeing during difficult times.

> *‘I’ve had a couple of other friends who said, look, don’t sit alone at home being miserable. If you need anything, if you just need to talk to someone, just give me a bell, or message me or whatever. So, that’s been nice, I think a lot of people didn’t realise how ill I was, and I think that when I went back to work, they were quite shocked, and were keen to reach out*.*’* (560_female_aged_50-54)

In the initial stages of illness, participants described receiving emotional and practical support from family and friends: ‘*<My sibling> used to do the shopping, and she’d sometimes do the cooking and bring it over*.*’* For some participants however, as their illness progressed and symptoms remained, emotional support became less available due to a lack of understanding or unwillingness to acknowledge that participants could still be unwell.

> *‘A lot of people don’t understand long Covid, so when you explain to them, I’m still not feeling right six months down the line, a lot of people have said, I think you’re just worrying too much. That’s what I think my parents come back with. They keep saying to me, you worry too much, there’s nothing wrong with you*.*’* (555_male_aged_45-49)

#### Peer support

Many were members of online long Covid peer support groups and found them empathetic and supportive: ‘*I was just feeling really validated that I’m not alone*.*’* These groups also facilitated access to information and resources on management strategies and services.

> *‘And the best support I got was from all the Facebook groups, believe it or not. That’s where I found a lot of information, because everyone else was on a similar timeline to me. So, we were all going through the same symptoms, so I knew I wasn’t going crazy*.*’* (552_female_aged_50-54)

Others reported mixed experiences of accessing online peer support groups, with some feeling overwhelmed *‘I think that can be quite scary when you listen to some of that’* and others acknowledging that some of the information was unhelpful or untrustworthy

> *‘I went on Facebook to go on one of those long COVID groups, which I looked at for a couple of days, but to be honest it started doing more harm than good looking at those, especially in the state I was in at the time*.*’* (566_male_aged_40-44)

On balance however, most participants found the groups helpful and actively filtered out the ‘*rubbish’*.

> *‘So, at least I feel like they’re trying to do something. There’s a lot on there, which is negative. I try not to read that because*…*But that’s like any social media isn’t it, you just have to choose*.*’* (553_female_aged_50-54)

### Theme 2: Access to services and treatments

While just over half of participants had received investigative tests, many participants described a lack of follow up ‘*we’re now on week 15 and I’ve still not had any results’* and ongoing stress and worry that the treatments or services they needed to manage their symptoms were unavailable, inaccessible or indeed were available, but not being offered to them.

> *‘What’s bothering me at the moment is I’m not in any plan or regime. So, I’m not on any pharmacological medicine. I’m not on any physical rehabilitation. So, I’m not on any programme. So, I just feel like I’m ambling. I’m just in the ether… A year of my life has gone. I can’t carry on*.*’* (553_female_aged_50-54)

Several participants described their frustration at being unable to access treatments or services because diagnostic tests were inconclusive or produced negative results, despite ongoing symptoms.

> *‘I had several chest x-rays, and things, so I was looked after. But there was nothing wrong as far as they could see, but obviously, I was still having symptoms. But I was a bit frustrated. It was several months by then, and I was thinking why has nothing been set up yet for people who are suffering?’* (552_female_aged_50-54)

While many participants described being aware of long Covid clinics, there was confusion around the referral process, ‘*we’d been rejected because they’ve changed the criteria’*, or there was a lack of available services in different areas of the UK.

> *‘There are no COVID clinics or anything in this area, so no. I have spoken to the doctors quite a lot since I’ve been ill, and they’ve been marvellous, but they’re still, we don’t know, we’re still waiting for the clinic to open*.*’* (561_female_aged_50-54)

Just under a third of participants who described being unable to access National Health Service (NHS) care had gone on to access private healthcare, either by paying for care themselves: *‘the only door that is half-open, is if I decide to pay and go privately’* or through insurance provided by their employer.

> *‘So, without that private healthcare, I don’t think I would maybe have got over some of the symptoms that I had because, as I say, the GP was just obviously it’s new for them as well, but they just weren’t offering any kind of help at all. And I remember when I went through them and says, well, I can’t actually refer you to the specialists with what you’re wanting under the NHS*.*’* (564_female_aged_70-74)

### Theme 3: Severe disruption to daily life

Most participants described leading active lives prior to COVID-19 infection, working in full time employment, participating in regular exercise and having busy social lives, however ongoing symptoms had significantly curtailed activities of daily living.

#### Daily activities

Ongoing fatigue and ‘brain fog’ were commonly described as having a huge impact on people’s working lives, with almost half the sample not able to fully return to work ‘*I couldn’t have even picked the laptop up and opened it, to be honest, you’re completely just wiped out*.*’* The impact on daily life for some participants was profound, resulting in difficulties *‘even being able to manage basic household chores’* or struggling to maintain personal care.

> *‘I haven’t showered for three days. I have to think about showering, I have to sit down and spend like two hours thinking about will I be able to manage showering today? Because even taking a bath makes me tired*.*’* (550_female_aged_25-29)

Some participants described experiencing a prolonged loss or change in taste and smell which resulted in no longer being able to enjoy food. For those who had previously used cooking as way to de-stress, this was experienced as particularly detrimental to wellbeing.

> *‘That’s kind of disheartening… I could make a really nice meal which before lockdown would’ve been the kind of thing that I would a) like to do, but also for me personally, I see it as more of if you’ve had a bad day I’m going to do a nice thing for myself and invest in making a nice dinner for maybe me and someone else*.*’* (565_male_aged_25-29)

#### Social life

Many participants were no longer able to participate in social activities, which had a profound impact on their mental health and wellbeing

> *‘I want to be able to have laughter, I want to be able to go out and be the life and soul of the party, which I’m just not anymore, so I do grieve*.*’* (557_female_aged_35-39)

For some, the severity of their ongoing symptoms meant they were unable to socialise, mainly because of fatigue and the need to conserve energy by pacing oneself ‘*I haven’t got the energy to interact*.*’* For one participant, socialising became difficult due to breathlessness which made holding a conversation difficult.

> *‘I couldn’t really talk to people… I just limited my <family member> to twice a week calls and I had to have short, five-minute calls because more than that then I’d have difficulty breathing. So that was a bit frustrating that I couldn’t actually talk to people*.*’* (569_male_aged_55-59)

Participants also described withdrawing from their social lives due to ‘*not wanting to worry people’* and feelings of ‘*guilt because I feel like I’m pouring more pressure on <my family>‘* by sharing their experiences with others. Others withdrew because they felt that they had little to contribute to friendships due to being unable to focus on anything other than their health *‘I was reflecting and thinking maybe I was talking about it all the time*.’

> *‘Well, early-days, I’d get emails and stuff and then it’s just drifted off. It’s my own fault because I could phone up and chat to people, but I just feel it’s a bit boring. How are you? You tell them how you are… How’s your heart? You tell them, so, I’d rather not dwell on it. And I’ve got nothing else to talk to them about now*.*’* (564_female_aged_70-74)

#### Enjoyable activities

Participants described the importance of engaging in new hobbies and activities or adapting previous activities to give them respite from their health problems, to support their wellbeing and to create a sense of achievement. Just under half of participants described engaging in low grade physical activity such as walking ‘*I’m just doing some very gentle, gentle stuff. I’m walking’*. Playing or listening to music or podcasts were important for distraction and relaxation.

> *‘When my relapse started and into three weeks of it I think it was when I was in bed most of the time, I had these awful, awful feelings of almost… I felt like crying but I wouldn’t, but I’d change my mindset to, oh I’ll do something more positive than lying in bed. I had some pastels and I was doing a little bit of drawing, and I’d listen to the radio and put music on and stuff. I’d just alter it just by changing my mind from focusing on, oh I feel ill and will I ever work again, will I ever be able to do anything again, to oh well I’ll do this*.*’* (559_female_aged_45-49)

For many participants however, participating in enjoyable activities that could improve or protect mental health was not possible, such as physical activity *‘I can’t do my exercises, and I can’t keep my body fit’*, getting out of the house ‘*I’ve been stuck at home for the whole year’*, or engaging in arts and culture.

> *‘I’ve been trying to do some interesting things, so I watched a ballet online from the Royal Opera House, I went to a book launch of a book. I thought, I’ve never done that before, I’ll do that online. All sorts of opportunities. But even the things that are joyful and enriching, and would normally be lovely and powerful, if I get really excited about it, it just makes me tired*.’ (563_female_aged_60-64)

Some participants described replacing their usual physical activities with competitive activities that could be undertaken at home such as online gaming.

> ‘*That (online Bridge) has been an absolute lifesaver for me because it’s kept my brain ticking over a bit, and given me, just a challenge, so I’m very fortunate with that. I’m sure other people have other things, but because I’ve got, naturally a competitive nature, with my golf, and I used to play tennis*.*’* (564_female_aged_70-74)

### Theme 4: Uncertainty of illness trajectories

Most participants described feelings of anxiety around the uncertain symptom trajectories they were experiencing, with fluctuating periods of having *‘good days’* of feeling well and even, at times, *‘euphoric’* followed by *‘bad days’*; periods of intense exhaustion and being unable to get out of bed. One participant described *‘the ebb and flow of it all’* as the most difficult thing to deal with.

> *‘There’s a lot of relapses involved… which, initially, was very, very demoralising, because you think, or I would think, I’ve turned a corner, I’m going to get better, and then it would just suddenly get, like take two steps forward and three or four steps back. And that’s characterised the entire journey I’ve had with COVID, up until now really*.*’* (560_female_aged_50-54)

For others, uncertainty about the future and how long their symptoms might continue led to increased anxiety about employment, finances, and family life. Participants described feeling less in control of their lives as time went on.

> *‘I can’t live like this, how am I going to go on in the future, am I going to get worse, am I going to get better? Obviously, you think of everything. How are you going to look after your kids, the family, your work situation, how are you going to pay your bills*.*’* (555_male_aged_45-49)

Participants also described having to weigh up the potential consequences of participating in activities that protected their wellbeing but that ultimately would lead to a relapse, against the immediate short-term benefits to their mental health.

> *‘I take the opportunities I can. So, a friend’s been off work and she messaged me yesterday and said, weather’s nice… do you fancy a walk in the park? And I said oh great, yes, lovely, I’m free from here and here. Technically, that was too much in a day, and I knew it was too much and I paid for it later in the evening and this morning, but it was so nice to see her*.*’* (563_female_aged_60-64)

### Theme 5: Changes to identity

Participants described profound changes to their sense of self because of long Covid *‘I’m a different person now’* and described feeling a loss of certain attributes and self-confidence that had previously defined who they were ‘*I think also it’s stripped me of me*.*’* For some, this change manifested itself as a loss of interest in self-care, a decrease in physical fitness ‘*I feel weaker’* and consequently changes to their physical appearance.

> *‘I don’t care anymore that I’m getting a bit soft in my body, that I used to do eight daily exercises and stuff like that. I don’t care that I’m looking older… and I suppose in some ways, that bothers me. Every now and then, I just think, this isn’t you*.*’* (564_female_aged_70-74)

> *I look like a big six-foot two bloke, but actually have jelly in my arms, just absolutely no energy in there, no physical strength at all. So yes, you feel less confident*. (566_male_aged_40-42)

Others described their identity as diminished because they were no longer able to fulfil their usual caring responsibilities and social roles as professionals, partners, parents and friends resulting in a sense of loss and feeling ‘*like a burden* or *waste of space*.’

> *‘I do feel a little bit lost because I just don’t know. I was a carer and now I’m being cared for. I’ve had to change, and I’ve struggled coming to terms with it a little bit*.*’* (561_female_aged_50-54)

> *‘So yes, there’s been a big change of identity. I’m a recovering patient from COVID rather than a meritorious consultant…’* (568_male_aged_60-64)

## Discussion

Our study found that people with long Covid experience unique challenges to their mental health and wellbeing related to the impact of symptoms on their lives, as well as interactions with health services, family and friends. Limited care and understanding from others, a lack of service and treatment options; disruptions to daily life and activities, the uncertainty of illness trajectories and changes to identity all contributed to a deterioration in mental health. Participants often described barriers to engaging in activities that might protect their mental health, however being listened to and feeling validated by health professionals, family and friends, accessing peer support and engaging in enjoyable activities within the limitations of their condition, were identified as important for psychological wellbeing.

Our findings align with, and extend, existing research into experiences of long Covid. They echo work that has previously described symptoms; impacts on daily life; and interaction with healthcare providers^16 28 29^. However, our work extends previous findings by highlighting which features of long Covid are seen as most significant in shaping mental health and wellbeing from the perspective of people with lived experience. Our findings also improve understanding of how these features are experienced by people with long Covid, and as such, present potentially useful considerations for healthcare, self-management, and wider society.

In keeping with the growing body of research, our participants reported multiple, varied, relapsing-remitting symptoms often severely impacting their ability to engage in daily life^16 28 29^. These limitations have strong negative impacts on mental health and wellbeing through limiting participation in enjoyable aspects of life and those that gave individuals their sense of identity, including work, leisure, and social activities. Experiences of loss and threatened identity were also found in a qualitative study involving 114 people with long Covid^29^, and research regarding other chronic diseases such as chronic fatigue syndrome^30^. Our findings identified the relevance of relapsing-remitting disease trajectories as important drivers of negative mental states, due to uncertainty about the future, which is well established as a driver of anxiety and depression for people with various chronic diseases^31^.

Other research, particularly from earlier on in the pandemic, also identified various barriers to healthcare^1 16 28 29^, including a lack of specialist service provision, and limited understanding of the illness from healthcare professionals. Our study also shows that some people resorted to accessing private healthcare and felt better supported to manage their symptoms. This finding raises concerns around health inequalities for people with long Covid who are unable to afford private healthcare. Potentially, with the creation of specialist long Covid services, increasing research, and clinical guidelines, such barriers should become less impactful over time, though given other competing needs faced by healthcare services, access could remain an issue. Feeling ignored or not believed by healthcare professionals, family members and friends, has been commonly reported^1 16 28 29^. These experiences are similar to those described by people with CFS/ME^32^. Our findings suggest such experiences have particularly powerful negative impacts on mental health.

Factors positively impacting mental health and wellbeing were infrequently mentioned. As in previous work, we did however find that interactions where people felt listened to and believed were greatly valued, and empathy from health professionals even when faced with limited treatment and referral options^12^ was important. Additionally, long Covid peer support groups appear valued by some individuals, though the potential for overwhelming and inaccurate information, as reported by participants in our study, is an important consideration.

### Strengths and limitations

Certain limitations should be discussed. Firstly, our sample included people who met clinical diagnostic criteria but without laboratory confirmation of COVID-19. This is in keeping with relevant guidance and excluding people without laboratory confirmation would have limited our ability to capture experiences of long Covid symptoms lasting up to 12 months as testing was limited in the first part of the pandemic. Secondly, a degree of selection bias may have occurred towards those with mental health and wellbeing impacts. Thirdly, our sample consisted of 21 people living in the UK, therefore assessing the external validity beyond this context is not possible, but likely to be relevant. Finally, online methods of recruitment and data collection may have excluded some participants without access to the internet from taking part, although this method did enable participants to be interviewed in their own homes across different parts of the UK.

### Implications

Our findings emphasise the need for accessible, patient-centred, specialist multidisciplinary healthcare provision. From healthcare professionals to family members and friends, ensuring people with long Covid are listened to, and their experiences validated, is vital. Importantly, optimising these experiences now features in learning modules and guidance for GPs^33^. Our findings regarding challenges in communicating long Covid experiences to family and friends suggest that it could be of value to adapt similar education and support materials and guidance for informal support networks. Given participants described specific mental health components of long Covid, these should be acknowledged as part of long Covid treatment pathways and potential contributory factors identified and addressed where possible. Supporting individuals with long Covid in adapting their work, social and physical activities to meet their health capabilities could also be valuable as part of tailored occupational health programmes. Long covid guidelines now include the ‘Pace, Plan and Prioritise’ principles around ‘pacing’ physical activity for self-management^34^, but whether this strategy improves symptom management remains unclear, and participants in previous research describe receiving conflicting advice^35^. Additionally, impacts on social participation suggests there may be a role for tailored community groups and social prescribing, though this should be accompanied by research and evaluation to assess appropriateness and effectiveness.

## Data Availability

The data are not publicly available due to their containing information that could compromise the privacy of research participants.

## Acknowledgements

The authors would like to thank those people who gave up their time to take part and contribute to the study.

## Author contribution

All authors meet the criteria for authorship as recommended by the International Committee of Medical Journal Editors. AB and DF had the original idea for the study. All authors (AB, HA, DF, KEJP) were involved in designing the study. AB provided guidance on qualitative methods including design and analysis. AB conducted the interviews. HA led on the analysis with support from AB and KEJP. Preliminary findings were discussed with the other authors and refined. The first draft of the manuscript was written by AB, HA and KEJP. All authors (AB, AB, DF, KEJP) read, contributed to, and agreed on the final manuscript draft.

## Patient consent for publication

Not required.

## Ethics approval

The authors assert that all procedures contributing to this work comply with the ethical standards of the relevant national and institutional committees on human experimentation and with the Helsinki Declaration of 1975, as revised in 2008. All procedures involving human subjects/patients were approved by the UCL Ethics Committee (Project ID 14895/005).

