## Supplementary material for "*‘I had no life. I was only existing’*. Factors shaping the mental health and wellbeing of people experiencing long Covid: a qualitative study": S1 Appendix

### Screening questions

**Thank you for your interest in taking part in our study on Social Distancing during Covid-19 and the impact of COVID-19 on health and wellbeing**

We need to collect some additional information to understand whether or not you are eligible to take part in the interview

**COVID-19 diagnosis status**

- ☐ Positive swab test
- ☐ Negative swab test
- ☐ Tested positive for antibodies
- ☐ Tested negative for antibodies
- ☐ Not formally diagnosed but suspected
- ☐ Other, please specify__________________

**If you have not received a positive test for COVID-19 or antibodies, did you experience any of the following COVID-19 Symptoms (please tick all that apply)**

- ☐ A continuous cough
- ☐ A high temperature
- ☐ Loss or change to taste/smell

**Date that you were diagnosed or suspected that you had COVID-19**

**_ _ / _/ 2020**

**For how long after the onset of suspected/confirmed covid have you been experiencing symptoms?**

- ☐ Less than 3 weeks
- ☐ 3-6 weeks
- ☐ 6-9 weeks
- ☐ 9-12 weeks
- ☐ More than 12 weeks (please state_______________)

**Can you describe the long-term health problems that you are experiencing following confirmed or suspected COVID-19? (please tick all that apply)**

- ☐ A cough
- ☐ A high temperature/fever
- ☐ Loss or change to taste/smell
- ☐ Sore throat and difficulties swallowing
- ☐ Fatigue
- ☐ Muscle pains/weakness
- ☐ Inability to concentrate
- ☐ Memory lapses
- ☐ Headache
- ☐ Aches and pains
- ☐ Shortness of breath
- ☐ Diarrhoea/vomiting
- ☐ Sleep difficulties
- ☐ New onset/poor control of diabetes/hypertension
- ☐ Skin rashes
- ☐ Chest pains

- ☐ Changes in mood, anxiety or depression
- ☐ Palpitations
- ☐ Thromboembolic conditions
- ☐ Other, please specify___________________

**Many thanks again for your time**
