## Supplementary material for "*‘I had no life. I was only existing’*. Factors shaping the mental health and wellbeing of people experiencing long Covid: a qualitative study": S2 Appendix

**Draft topic guide: Adults with long term health problems following suspected/confirmed Covid**

**Ask to describe 'normal life'**

- Employed? Type of job, hours etc,
- Education/study
- Full time parent or carer?
- Use of any community services?
- Who you normally live with, does this change, separated/ extended family?
- If they have a long-term condition/cancer they mentioned (what condition, when diagnosed, if on or whether they have had complete treatment)
    - What was current treatment plan? How was it being managed? What was usual routine for appointments/follow-up?
- Whether you would usually have done any type(s) of regular exercise (whatever they perceive as exercise including walking/gardening)

**IMPACT OF SUSPECTED/CONFIRMED COVID**

**Ask to describe *onset* and *initial* impact of Covid-symptoms**
Prompts include:

- Can you tell me about when you first suspected you had Covid-19? (symptoms, how did you feel, how did it affect you, under what circumstances do you think you contracted it (e.g. work, socialising, from someone in your household)
- Were you able to get tested? (why/why not, explore facilitators and barriers to testing, how did you feel about the outcome?)
- What was the impact on "normal" life at the time? (employment, education, caring responsibilities, living situation, impact on other health conditions, ability to exercise/participate in healthy lifestyle behaviours)
- Were you able to access treatments, services, support (why/why not)?

**Ask to describe ongoing impact and development of longer term health problems**
Prompts include:

- Can you describe how your health has been impacted?
- What has been the impact on "normal" life? (employment, education, caring responsibilities, living situation, impact on other health conditions, ability to exercise/participate in healthy lifestyle behaviours)
- Have you been able to access any treatments, services or support (why/why not)?

LONG TERM CONDITION/CANCER (if applicable)
**How has Covid-19 had an impact on being able to manage the [long term condition/cancer]?**
Prompts include:

- What has been the impact on any normal appointments? (cancelled, delayed, unable to speak to appropriate healthcare professional, changed to different method/location of appointment e.g. online/telephone)
- What has been the impact on any treatment? (cancelled, delayed, changed from usual treatment plan, unable to get medication/prescriptions)
- Impact of suspected/confirmed Covid diagnosis on symptoms/access to treatments/services?

- Impact of longer term health problems from suspected/confirmed Covid on the health condition itself?
- Impact of longer term health problems from suspected/confirmed Covid on ability to access services and treatments for existing health condition?
- Have you experienced any other impact on any symptoms/side effects?
- How have you felt about [any mentioned changes/impact above]?

UNDERSTANDING AND ADHERENCE TO GUIDELINES

**Are you, or have you been self-isolating? (how long for, reasons for this) a key worker, working but not a key worker, social distancing/ 'staying at home'**

- Please describe what this is for you and your family/ household?
    - i.e. are you self-isolating with neighbours helping to get groceries, or going out for these?
    - Or self-isolating with no outside exercise? Etc – if self-isolating without outside exercise how are you feeling about this? Do you have a garden or outdoor space?

- **What do you understand by the 'social distancing' advice that is being given – what does it mean to you?** Have you been…
    - Avoiding crowds
    - Keeping personal distance from others
    - Isolating
    - Avoiding close contact greetings
    - Socialising/going out only with those in your household
    - <if exercising outside> are you finding places to go where you can keep your distance from others?

**Have you been able to stick to the social distancing advice that has been given to your group? Please tell us about why/ why not?**
**[COM-B prompts can be used here, to include:]**

- Impact of suspected/confirmed Covid
- Impact of longer term health problems from suspected/confirmedCOVID
- Any existing physical or mental health problems
- Group membership/ applicability
- Beliefs about consequences/ health beliefs
- Consequences for others/ self
- Needing to work/ living arrangements, whether others are self-distancing in the same house/area
- Work/ Caring responsibilities, providing emotional support
- Peer pressure to socialise
- Government rules/punishments <prompt to ask how they feel about the Government recommendations that are relevant to exercise for them>,
- Feelings about losing normal life
- Change of routine/ habits

**SOCIAL LIFE**
**How would you describe your social life beforeCovid-19?**

- How would you describe your social network – for example size, types of people, types of relationships, do they live with you, nearby or further away, how often do you see each other, how well do you know each other? How do you interact, face to face, online or social media?
- Social activities?
- Could you describe any community services/participation or volunteering participation?

- Could you describe the social support you have? (such as emotional support, advice and information, someone to help you with money or milk/bread/essentials, community services)
- Can you tell us about any ways your social networks/ friendship groups influence you, such as peer pressure, or encouraging you to get involved in things? Do you compare your life to theirs?
- Social engagement (social roles, bonding, attachment)

**How would you describe your social life during the pandemic? Please tell us about this**
Prompts include:
- Has your experience of Covid/long Covid had any impact on your social life? (explore all prompts below in relation to this)
- How would you describe your social network – for example size, types of people, types of relationships, do they live with you, nearby or further away, how often do you see each other, how well do you know each other? How do you interact, face to face, online or social media?
- Social activities?
- Could you describe any community services/participation or volunteering participation?
- Could you describe the social support you have? (such as emotional support, advice and information, someone to help you with money or milk/bread/essentials, getting medication/access to healthcare, community services)
- Can you tell us about any ways your social networks/ friendship groups influence you, such as peer pressure, or encouraging you to get involved in things? Do you compare your life to theirs?
- Social engagement (social roles, bonding, attachment)
- Any negative responses/changes in relationships with anyone in your social network as a result of long Covid?

MENTAL HEALTH
**How do you feel about the changes that have been brought about by Covid?**
**Have they had any impact on your mental health or wellbeing? Please tell us about these**
- What are the things most bothering you at the moment?
- Have you experienced any impact on positive emotions? (prompts: how deeply you can engage with what you are doing, sense of meaning/ purpose, relationships with others, how well you are managing and feelings of control over your situation?)
- Has there been any impact on your sense of identity?
- Have you experienced any stigma due to long Covid?
- Have you experienced any negative psychological feelings? (prompts: such as shame, guilt, lack of pleasure, anxiety, worry)
- Please tell us about any physical symptoms due to being stressed or anxious? (prompts: fatigue, sleep problems, pain, illness symptoms, palpitations)

**Have you been doing/ planning anything to help with this?**
- Connecting with family or friends/ work colleagues online?
- Online groups?
- Hobbies/ Reading
- Exercise at home <ask about what they have been doing and if there are specific resources they have found useful to exercise>
- Volunteering
- Other engagement

**Why are you doing/ not doing these things?**

- Long Covid? How/why?
- Helpful/ not helpful – please tell us why
- Enjoyable
- Good for mental health/ wellbeing
- Can't get online, not connected, not comfortable, affordability, confidence in using/ skills
- Skills in using the internet/ communication software
- Living arrangements/ Work/ caring demands
- Peer support/ pressure
- Difficulties/ restriction in physical environment

PROSPECTION

**Has any aspect of the pandemic/long Covid meant that you have any worries for the future?**

**How are these different from the worries you had before?**

- Sense of control/ powerlessness
- Severity of worries / perspective
-

**Will this change the way you live your life in future?**

- The way you connect with others
- How you look after yourself
- How you support others
- How you work?
- How you exercise?

Has this changed any of your priorities for the future?
